# Spatial scales in human movement between reservoirs of infection

**DOI:** 10.1101/2020.04.17.20069047

**Authors:** Robert J. Hardwick, Carolin Vegvari, Benjamin Collyer, James E. Truscott, Roy M. Anderson

## Abstract

The life cycle of parasitic organisms that are the cause of much morbidity in humans often depend on reservoirs of infection for transmission into their hosts. Understanding the daily, monthly and yearly movement patterns of individuals between reservoirs is therefore of great importance to implementers of control policies seeking to eliminate various parasitic diseases as a public health problem. This is due to the fact that the underlying spatial extent of the reservoir of infection, which drives transmission, can be strongly affected by inputs from external sources, i.e., individuals who are not spatially attributed to the region defined by the reservoir itself can still migrate and contribute to it. In order to study the importance of these effects, we build and examine a novel theoretical model of human movement between spatially-distributed focal points for infection clustered into regions defined as ‘reservoirs of infection’. Using our model, we vary the spatial scale of human moment defined around focal points and explicitly calculate how varying this definition can influence the temporal stability of the effective transmission dynamics — an effect which should strongly influence how control measures, e.g., mass drug administration (MDA), define evaluation units (EUs). Considering the helminth parasites as our main example, by varying the spatial scale of human movement, we demonstrate that a critical scale exists around infectious focal points at which the migration rate into their associated reservoir can be neglected for practical purposes. This scale varies by species and geographic region, but is generalisable as a concept to infectious reservoirs of varying spatial extents and shapes. Our model is designed to be applicable to a very general pattern of infectious disease transmission modified by the migration of infected individuals between clustered communities. In particular, it may be readily used to study the spatial structure of hosts for macroparasites with temporally stationary distributions of infectious focal point locations over the timescales of interest, which is viable for the soil-transmitted helminths and schistosomes. Additional developments will be necessary to consider diseases with moving reservoirs, such as vector-born filarial worm diseases.

## 1. Introduction and background

Defining the spatial scales over which transmission should be considered is a relatively recent research area in the context of neglected tropical diseases (NTDs), where much of the effort has been focused on the development of geostatistical methods for each disease in turn [1, 2, 3, 4, 5, 6, 7, 8, 9, 10, 11]. In particular, mathematical models of helminth transmission have only recently begun to incorporate the dynamical effects of human movement between reservoirs of infection [9, 12, 13, 14].

Helminth infections (or helminthiases) are a class of macroparasitic diseases which include, among others, the soiltransmitted helminth (STH) infections, schistosomiasis and lymphatic filariasis (LF). Following World Health Organisation (WHO) guidelines, free drugs have been donated by pharmaceutical companies since 2010 [15, 16, 17, 18, 19] to countries significantly affected by helminth-associated morbidity. Control initiatives have been developed specifically to investigate the prospects for elimination as a public health problem in the long term [20, 21].

In Fig. 1 we have illustrated the essential role that infectious focal points have on driving the transmission of human helminth infections. In the case of STHs, these focal points consistute eggs, or larval stages in the soil, that are either ingested or enter the body via skin penetration. Similarly, in the case of schistosomiasis transmission, these focal points exist in water sources where larvae, that are released by freshwater snails (intermediate hosts), penetrate the skin to infect human hosts.

**Figure 1.**
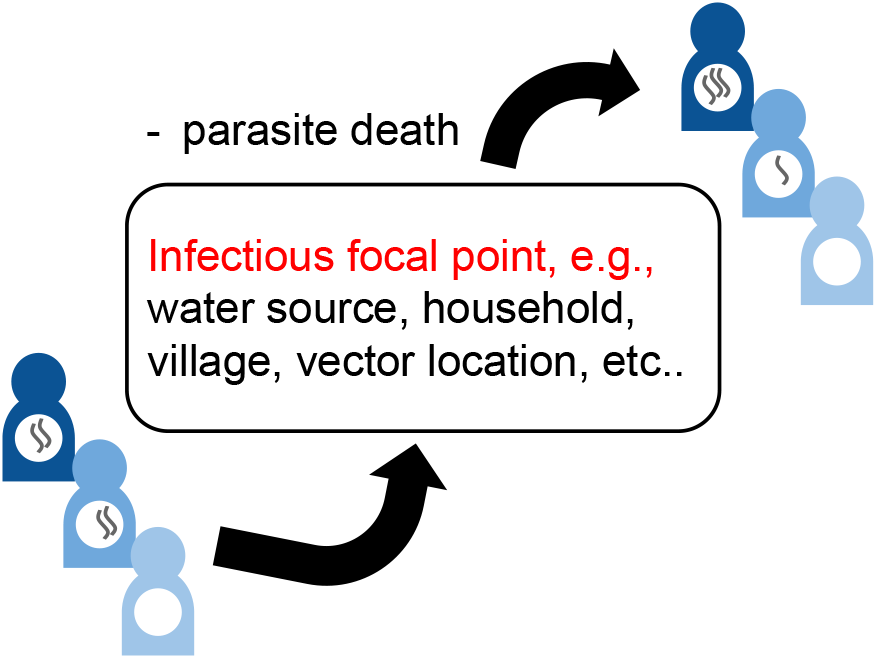
Illustration of the transmission dynamics of human helminth infections via a defined focal point.

It may be possible to minimise the effect human movement has on evaluation units (EUs) of, e.g., mass drug administration (MDA) control programmes, by specifying a critical spatial scale around the focal points within a reservoir (and corresponding to a human community) over which one may sample and treat infections. At this scale (or larger), the number of migrants contributing to the reservoir per unit time is potentially low enough such that their effect may essentially be neglected. If such a scale exists it would provide a natural level of res-olution for those generating spatial maps of the prevalence of infections in a given region. Note that for some NTDs, implementation units (IUs) and evaluation units (EUs) differ. In this paper, we shall look at the spatial scale of EUs because there is currently no evidence-based framework for choosing an appropriate spatial scale at which programme managers can evaluate the impact of NTD programmes. We will demonstrate the existence of this scale, which is expected to vary between helminths and regions, by developing a spatial model of human movement between defined locations which is consistent with the current literature.

The distances travelled by human movement patterns are not purely random, but exhibit certain known characteristics similar to those of known random walks, e.g., the same heavy-tailed distributions as those of Lévy flights [22, 23]. Furthermore, in a variety of countries and levels of urbanisation, Ref. [24] uses mobile phone data in many different countries to conclude that there is evidence for some universality in the distribution shape of daily work-home commute distances, which appear to fit a broken power-law. Note that the distributions are similar to, e.g., those observed for radial distance in Refs. [25, 26], where these works make use of geolocated tweets and GPS data, respectively. The findings in all cases above, and other examples of broken power-laws for human mobility in the NTD disease modelling literature (see, e.g., Refs. [27, 28]), will motivate us to use a similar description to develop a simple model of human movement from households to focal points of infection in this work.

In Sec. 2, we derive a broken power-law model with one jump per individual, which both replicates the distribution behaviour and also suggests a possible mechanism for its origin. The model we introduce, however, will remain flexible (by varying parameters) to other possible power-law descriptions for human mobility and so we will not be wholly reliant on one possible description. In Sec. 3 we briefly investigate a computationally-efficient extension to this model which uses an approximation to describe multiple jumps per individual for movement patterns with longer timescales — both of which might be important for an accurate representation of the rele-vant human behaviour.

Having developed our model and illustrated its possible extensions, in Sec. 4 we apply it to obtain a critical spatial scale at which the migration of individuals to focal points clustered as infectious reservoirs declines to a negligible level. This result is shown to vary with helminth species, human movement patterns and regional geometry of settlement patterns. The analysis also provides insight into the most important pieces of (often missing) information which are necessary to build an accurate model of human migration patterns between reservoirs of infection. Collecting such information will be key to the success of future helminth control programmes in reaching their targets, e.g., achieving STH elimination as a public health problem [29]. Lastly, in Sec. 5, we discuss how the spatial extent of reservoirs of infection considered here should influence the scale of EUs for control programmes and conclude with a summary of our findings and prospects for future work.

## 2. A one-jump model

### 2.1. Locations of infectious focal points and households

There are important ethical implications to the public availability of data on both the spatial locations and movement patterns of individuals over various scales in time, especially in low-and middle-income countries [30]. Among these are data privacy and the potential identifiability of individuals through the use of ‘big data’ by governments, companies and universities, and the potential use of this information for non-research purposes. Therefore, access to such high-quality data for study is scarce. This represents a challenge for those concerned with the various consequences of such movements and their methods of model validation. In this paper, we present mathematical models that can be used in the absence of access to high-quality movement data. Model dynamics are implemented on the spatial patterns of, e.g., building locations, obtained from real-world datasets, e.g., the high resolution settlement layer dataset generated by the Facebook Connectivity Lab [31]. Our methodology should allow researchers to draw useful conclusions on the impact of human spatial movement on NTD programmes from readily available geospatial data (coordinates of households, settlements and environmental features, such as water bodies). Other data sources, e.g., call data records (from mobile phones) and data from migration questionnaires in control studies, may be available under certain circumstances and can easily be integrated in our modelling framework. The models we introduce here should also be able to take this specific case-study information into account by appropriate parameter inference.

Let us first consider the embedding of spatial locations of households as points in a 2-dimensional flat space. In addition to these household points there will also exist focal points of infectious contact that depend on the helminth species, e.g., near buildings, water sources, parts of farmland with large concentrations of infectious material and households themselves. Choosing a particular focal point, we will use the observed spatial distributions of buildings in the high resolution settlement layer dataset [31] as a proxy for the spatial distribution of household locations around it. Therefore, the expectation of having *n*_*<r*_ nearby households or buildings within a radial distance of *r* from the focal point, i.e., E(*n*_*<r*_), scales according to the following radially-dependent power law

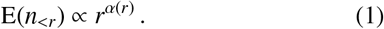

It is important to note that, since at first we consider only a single focal point from which household distances is measured, the distribution of focal points themselves is not relevant to the calculation. The spatial distribution of focal points will, however, become relevant later on when we cluster them into reservoirs of infection and investigate their spatial extent. Note, however, that households themselves may be a reasonable tracer of the underlying distribution for infectious focal points of STH [32, 33], though our calculations throughout will not depend on this possibility.

In the cases of STH and schistosomiasis transmission, although it is clear that the location and importance of their infectious focal points can vary over time — e.g., a chance decline in snail population for a particular location at the edge of a fresh water source leads to a drop in infectious spread — throughout this work, we shall assume that their spatial distribution is effectively temporally stationary over the timescales of interest. Note that for some NTDs, the infectious contact event locations can also be mobile, e.g., in the case of lymphatic filariasis transmission, where the larval stage of microfilariae enter the body by bites from infected mosquitoes — hence, the contact event locations will migrate with the human and mosquito populations.

By averaging over the observed point spread patterns in the high resolution settlement layer dataset, we obtain the following cumulative neighbouring point number distributions as a function of radial distance. This can be compared to the radial dependence of the power-law index *α*(*r*) we have quoted in Eq. (1). In order to give an indication of the effect of spatial heterogeneity in the distances between buildings for this averaged quantity, we have plotted some example lines (left column of plots) and the sample mean lines with root-mean-square deviation (RMSD) shaded regions of *α*(*r*) (right column of plots) from individual randomly sampled initial building locations for different countries in Fig. 2. In Central Malawi and Northern Benin, up to some substantial variance, below a transitioning radial scale *r < r*_tr_, the index seems to have a mean of *α ≃*1 and above this scale *r > r*_tr_ the mean of the index begins a transition to *α* → 2. In the Central Ivory Coast, we see that the mean trend rapidly varies between *α ≃* 1.5 and 0.5 below the transition scale *r < r*_tr_, and then at radial distances much greater than this scale *r*» *r*_tr_ most of the power-laws appear to begin a transition to *α* →2. The substantial variation below the transition scale in the Ivory Coast is likely due to a greater degree of clustering on particularly small scales (note the drop towards *α ≃* 0 before the transition scale indicates this as very few new points are added to the cumulative total for this range).

**Figure 2.**
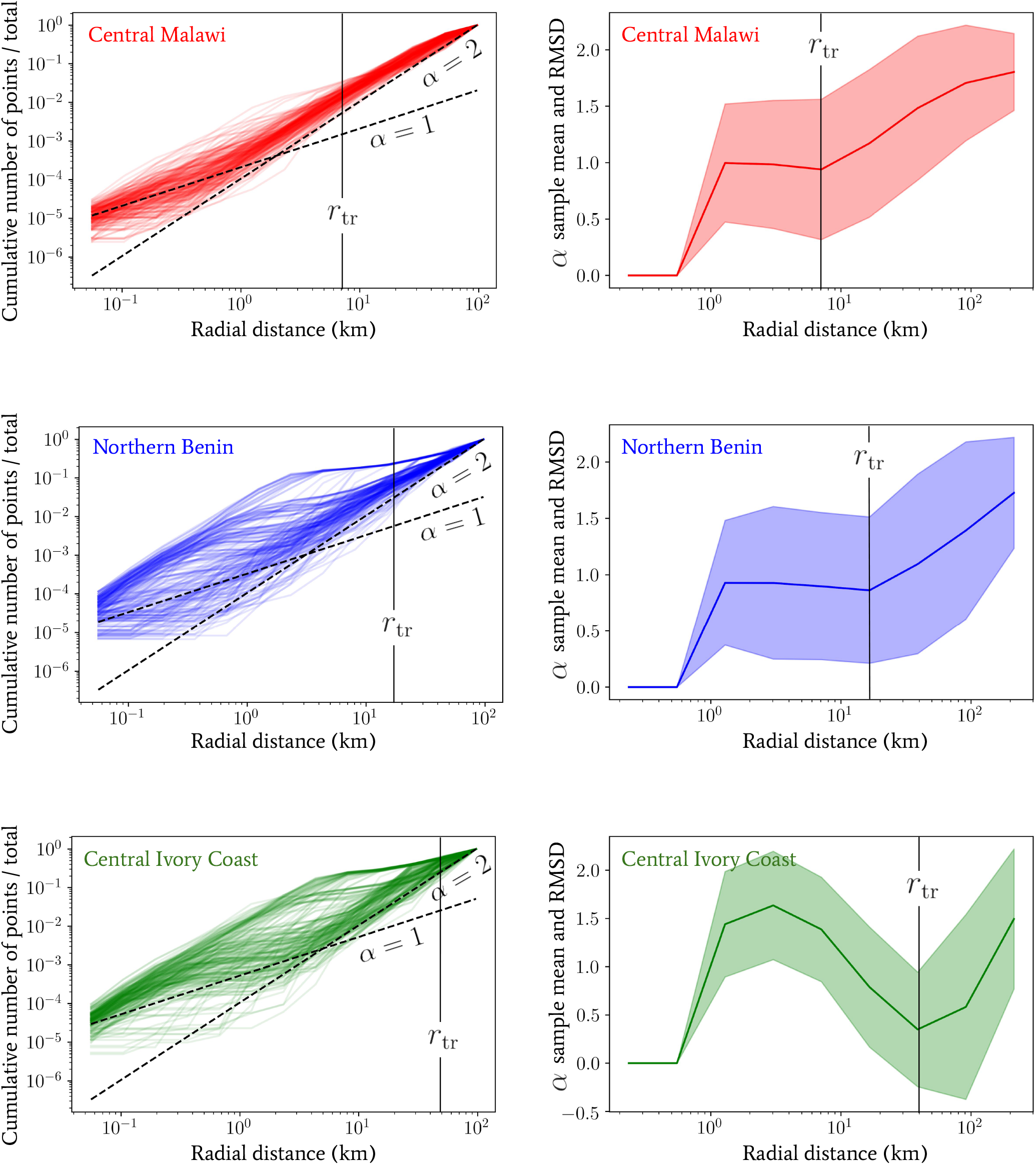
*Left column:* The cumulative number of buildings (vertical axis) below a given radial distance (horizontal axis) from sample locations in Malawi, Benin and the Ivory Coast obtained from the high resolution settlement layer dataset generated by the Facebook Connectivity Lab [31]. Lines for different constant *α* and *r*_*µ*_ values are provided only as rough indicators for comparison. *Right column:* The sample mean lines and root-mean-square deviation (RMSD) shaded regions of the power law index *α* plotted against radial distance in each case. The RMSD was calculated by computing discrete derivatives between log-frequency bins as a function of log-radial distance. In each country, the collection of sample locations was randomly drawn from: the central region of Malawi, including Lilongwe; the wide northern region of Benin, spanning between Kandi and Djougou; and a wide central region of the Ivory Coast, including Yamoussoukro.

For all countries and initial locations in Fig. 2, we can confirm that the majority of the power-laws begin to converge to a mean of *α* → 2 above a particular transition scale *r*_tr_ which varies substantially between location. This convergence reflects the emergence of statistical spatial homogeneity only on very large spatial scales, but this is clearly not present below *r*_tr_ in all cases. The power-law signature of statistical homogeneity being *α* = 2 should become clear by noting that another way to generate it is to draw a Poisson point process with an intensity *r*d*r ∝ r*^2^ (the flat Euclidean measure in 2 dimensions). Note that statistical isotropy is also implicitly confirmed by choosing different *r* = 0 locations for each line and observing the same behaviour up to some noise that is captured by the RMSD in the right column of plots. Deviations from this trend towards *α* = 2 (particularly in the Ivory Coast) are also present in many other locations in the same countries, due to the presence of substantial global heterogeneity, e.g., through proximity to large voids in the distribution of points, which exist for geographic reasons.

### 2.2. Obtaining a model using maximum entropy arguments

In this section we provide a simple argument to generate the broken power-law frequency distribution of work-home commute distances (as observed and described in, e.g., Kung et al. [24] and Zhao et al. [26]) using maximum entropy arguments. The distributions of movement distances we derive will be used to model the migration of individuals from their household locations to a focal point of infection. In the absence of access high-quality site-specific data that we discussed in the previous section, we are using the work-home commuter distance distributions as a guide for the likely patterns of movement. Work-home commuter distances should be a reasonable proxy for the overall movement of individuals in many areas. However, the model that we develop in this section is adaptive and flexible to modification, even in circumstances where this proxy is inappropriate, we will be able to vary distribution parameters to quantify their impact on our conclusions.

For greater analytic insight in the expressions we derive in this section, and throughout most of the subsequent arguments made in this article, we shall now assume that Eq. (1) has a constant index *α*(*r*) = *α*. When considering the heterogeneity in the real power-law behaviour below *r*_tr_ in Fig. 2, this assumption will not be appropriate in all locations/situations. Note, however, the results we obtain in all cases may be easily numerically generalised to take into account complex transitions between different power-law behaviours, as are observed in Fig. 2. In addition, choosing several key values for constant *α* (which fit within the range of observed power law behaviours) in what follows will aid with quantifying its impact on our conclusions.

Assuming that each individual jumps to a single new location and then returns back to their starting position over the course of each day, we may follow a simple argument that incidentally generates a distribution over travel distances which is consistent with those observed and described in, e.g., Kung et al. [24] and Zhao et al. [26]. Let us take the distribution of neighbouring points to be isotropic. Defining *p*(*r*) as the probability density function (PDF) of an individual jumping to a given location as a function of its radial distance, one may then transform the flat radial probability measure *p*(*r*)d*r* by a Jaco-bian *v*(*r*) = dE (*n*_<*r*_) / d*r* like so

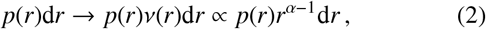

which accounts for the power-law density of points that may depart from linear scaling according to Eq. (1). Another way to view this rescaling is to consider the expectation of the probability measure for a radially-generated, statistically isotropic Poisson point process.

Note that if we additionally assume that the PDF of the jump distribution for each individual has a mean *σ* then the maximum entropy PDF is that of an exponential distribution *p*(*r*) = *p*(*r*; *σ*) = Exp(*r*; 1*/σ*), then by using Eq. (2) we can trivially demonstrate that the resulting normalised PDF is a Gamma distribution

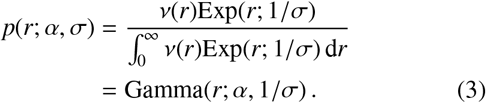

One possible interpretation for *σ* here is a number, with physical units of distance, which quantifies the access of that individual to varying degrees of transportation methods that are themselves capable of travelling varying distances. To clarify, without access to any motorised transportation, an individual may receive a low value for *σ*, and in contrast, if an indvidiual has access to a vehicle then they may receive a larger *σ* value. Despite this simple suggested explanation, the presence of variable scales for human travel distance distributions is not contingent on access to vehicular transportation at all. Indeed, human walking patterns themselves are known to exhibit multi-distance-scale characteristics in many contexts — see, e.g., Ref. [34].

Assuming that 1*/σ* values may vary across the population of individuals with a distribution of known mean E(1*/σ*) and expected scaling E(ln *σ*), the maximum entropy distribution for values of 1*/σ* is another gamma distribution. Choosing specific parameter values, we therefore have

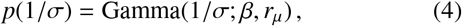

where the resulting marginal jump PDF is obtained by integration in the following way

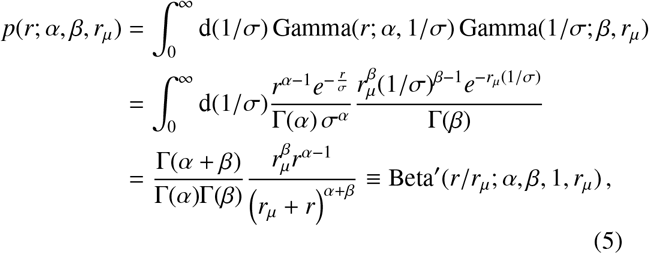

where Beta,(.;.,.,.) denotes the generalised beta prime distribution (or beta distribution of the second kind), which takes a form similar to that of a beta distribution. Note that, to avoid a divergent integral, one must specify that *α, β, r*_*µ*_ ∈ ℝ ^+^.

The main motivations for using maximum entropy probability distributions in the case above (and in general) include the beneficial properties that such distributions have when limited *a priori* information is available. In the present instance, these properties translate into distributions which account for the maximum amount of potential variability in outcomes while still satisfying the basic pre-defined classifying properties, e.g., the gamma distribution we have used above is maximally entropic under the condition that the mean and expected scaling of the distribution are known. Furthermore, it is often reasonable to expect that the steady states of many systems observed across the sciences are well-described by maximum entropy configurations.

We introduced another radial spatial scale *r*_*µ*_ (distinct from *r*_tr_ in the previous section) in Eq. (4) whose effect, in conjunction with *α* and *β*, we shall explore. In Fig. 3 we have plotted the marginal jump PDF given by Eq. (5) for a range of parameter values *α, β* and *r*_*µ*_. By inspection of Eq. (5), in the limit where *r* « *r*_*µ*_ the distribution scales as ∼ *r*^*α*−1^, whereas in the opposite limit *r* » *r*_*µ*_ the distribution exhibits a scaling ∼ *r*^−*β*−1^. The scale of *r*_*µ*_ therefore plays the role of separating two regimes in the distribution of jump lengths for a given individual. For radial distances below *r*_*µ*_, the geometric distribution of available spatial locations to jump to (which is encoded in the *α* parameter) dominates the behaviour, whereas, for radial distances above *r*_*µ*_, the predisposition of individuals to travel a given distance combines with the geometry of points to give the distribution behaviour and this is encoded in the *β* parameter of the second gamma distribution in Eq. (4).

**Figure 3.**
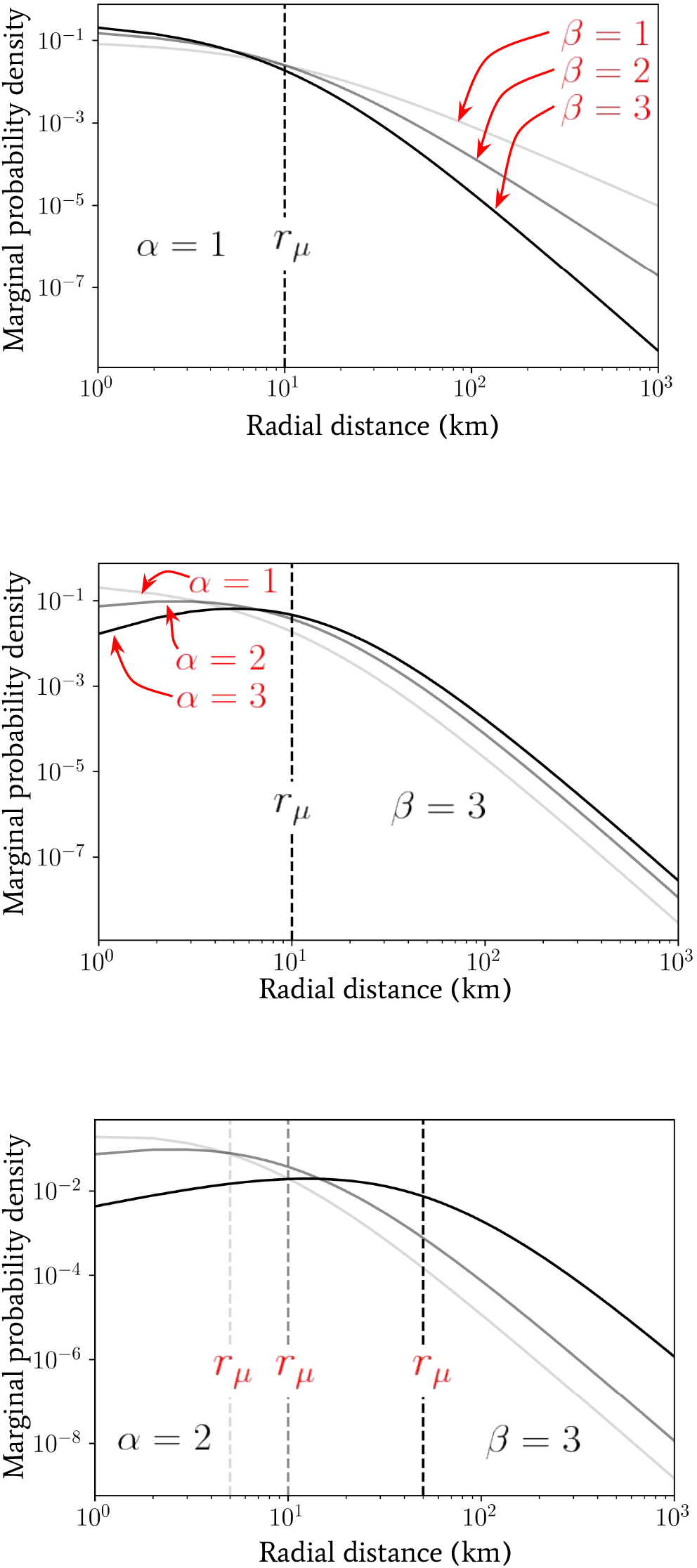
Plots of the marginal probability density *p*(*r*; *α, β, r*_*µ*_) as a function of radial distance (in units of km) generated using Eq. (5) for a range of *α, β* and *r*_*µ*_ values for comparison.

So far we have *not* assumed that the parameter *r*_*µ*_ in the distribution over inverse-jump scales 1*/σ* given by Eq. (4) is related to any purely geometrically-driven break in the power laws of Fig. 2. By comparing our observed power laws for the Ivory Coast with the observed work-home commuter distance distributions in Ref. [24], for instance, we see that *r*_*µ*_ « *r*_tr_ in general. This is perhaps unsurprising since the scale *r*_*µ*_ may arise due to the availability of easy access to vehicles that allow for longer-distance travel for either work or school, or water sources near a given settlement. Despite this fact, it is intriguing to note that the radial scale associated to the peak in clustering (*α* ≃ 1.5 well below *r*_tr_) for the Ivory Coast dataset in Fig. 2 occurs at the same rough scale that is observed for *r*_*µ*_ in Ref. [24]. We leave the investigation into the potential link between these two scales for future work.

Despite statistical isotropy and homogeneity on large spatial scales, we have observed that the variation between the power-laws for cumulative building numbers observed in Fig. 2, and hence the *α* index of Eq. (1), exhibits a large degree of location dependence between regions. In Fig. 4, we see the effect that this small-scale heterogeneity in the distances between buildings has on the probability density for our one-jump model. We plot a comparison between the binned frequency of 10^4^ individuals (with exponential jump distributions each with a scale drawn from Eq. (4)) simulated to travel (solid lines) between two buildings on real-world map data. The data for building locations corresponds to the same regions of each country as in Fig. 2, and the marginalised jump probability densities derived from Eq. (5) for choices of *α* = 1, 2. In all cases we have fixed *β* = 2 to a single value for easier comparison of different lines; this value is approximately the same as the tails observed in the distance distributions of Kung et al. [24].

**Figure 4.**
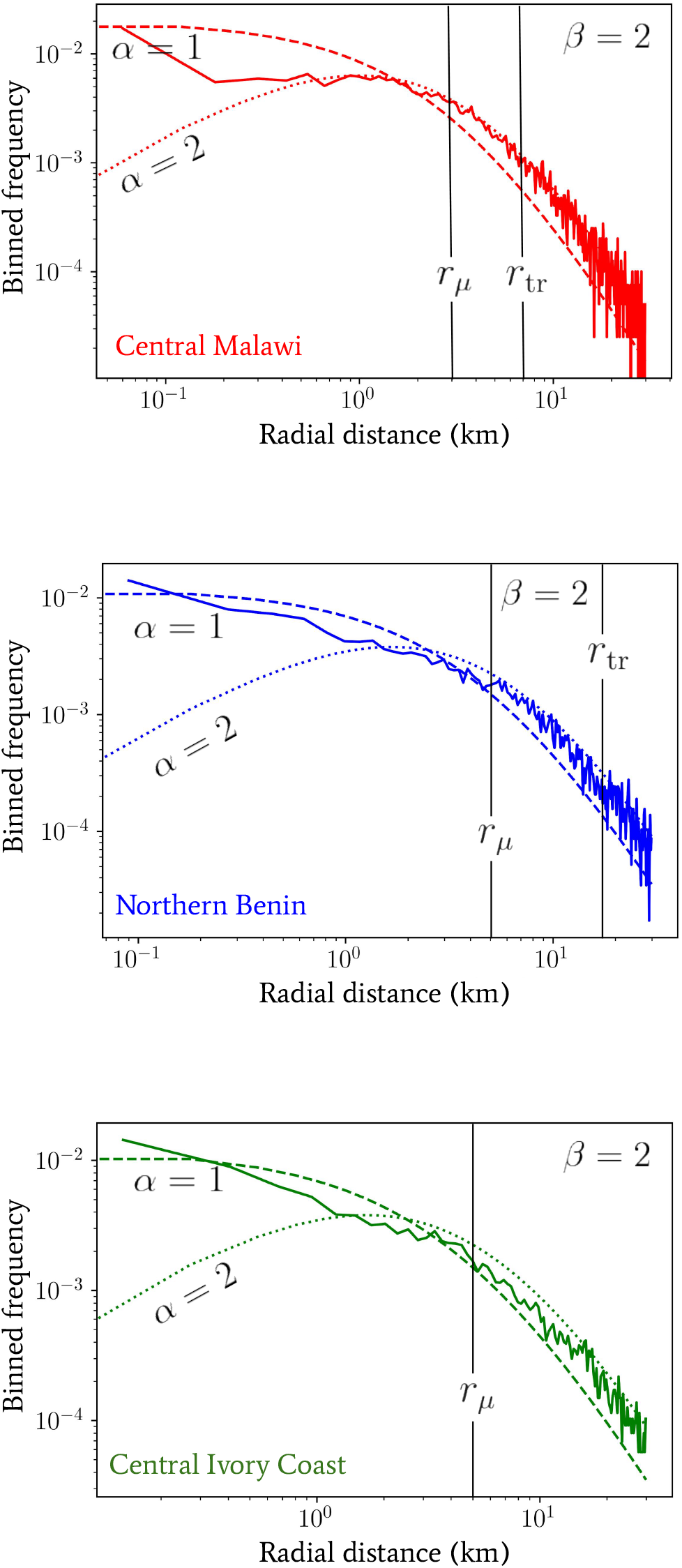
The binned frequency of 10^4^ individuals simulated to travel a between two buildings (solid lines) whose locations correspond to data from the same regions of each country as in Fig. 2 (where some globally-applied uniform random point density thinning has been used to reduce computational load of exploring the distribution tails). This corresponds to the same marginalised jump probability density as Eq. (5) but instead of using a geometric power-law given by Eq. (1) we have implemented our single jumps on real-world map data (with exponential jump distributions each with a scale drawn from Eq. (4)). For comparison, we have also plotted some jump probabilities calculated using Eq. (5) with *α* = 1 (dashed lines) and *α* = 2 (dotted lines). In all cases we have fixed *β* = 2.

The agreement between the *α* = 1, 2 curves and the distances generated from single simulated jumps between buildings on the real-world map data appears to be variable between countries. Overall, it is unsurprising that the shapes of the curves all seem to agree for scales *r > r*_*µ*_, because the asymptotic ∼ *r*^*β*−1^ behaviour dominates in all cases, which is consistent with our model. In the opposing limit, however, the small-scale variability in power laws for each country dominates and the departure from our constant-*α* models appears more pronounced — in favour of more transitory distance-dependent behaviour. Note also that the range in variability that the region between our *α* = 1 and *α* = 2 lines can account for is quite substantial. Such a range in behaviour motivates the use of both *α* = 1 and *α* = 2 in quoting the most important results that follow so as to correctly assess the impact of small-scale region-specific building distributions.

## 3. Multi-jump processes

In the previous section, a single jump model from a household to an infectious focal point location was developed in order to describe the movement of individuals between locations before contributing infectious material. An important caveat to this treatment of movement patterns is that one must also consider the possibility that multiple movements are performed either: between buildings before making a contribution to a focal point, or between infectious focal points themselves, supplying each some portion of infectious material in turn. The latter of these will be very difficult to quantify without some form of data, but for the former, we can explore some potential modifications to our model.

In this section, we will briefly explore an extension to the basic model of human movement presented in Sec. 2 to include successive jumps by an individual to multiple buildings before reaching an infectious focal point. The type of human movement we aim to capture is not just that of daily commuting, but also potentially longer distance travel across multiple days/weeks. The latter form of movement can contribute to the variability in migrant seasonal labour or family visits which can change the population numbers in a given region on a yearly timescale.

### 3.1. Homogeneous randomly-directed jumps

Following along similar lines to Sec. 2.2, let us now consider an individual who completes multiple successive jumps with exponentially-distributed lengths *r*_*i*_ (where we have indexed each successive jump event with an *i*) with a fixed mean scale *σ* within a day. For a fixed jump rate *𝒥* in time, consider the following compound Poisson process for a two-dimensional vector ***x***(*t*) which encodes the 2-dimensional Euclidean coordi-nate position of an individual over time

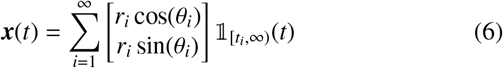

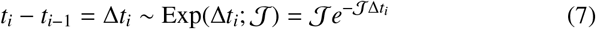

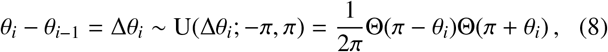

where U(Δ*θ*_*i*_; −*π, π*) is the uniform distribution PDF (assuming statistical isotropy) over the change in angle and Θ(·) is a Heaviside function. In Eq. (6) above, note that we are using an indicator function 𝟙_*A*_(*t*) which takes value unity if *t* ∈ *A*, else zero.

The process specified by Eq. (6) assumes isotropy of both the geometric distribution of the points and a uniform-random direction choice of the individual. The distribution *p*[***x***(*t*)] is not known as a closed-form expression, however, it clearly has both a vanishing first moment

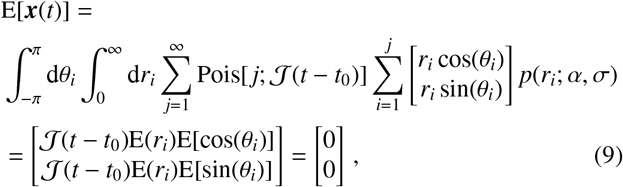

and a second moment which scales according to *σ*^2^

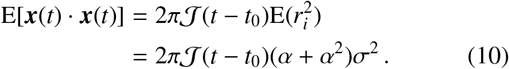

Furthermore, with a scale distribution over jump lengths applied — as in Eq. (5) — the second moment given in Eq. (10) becomes

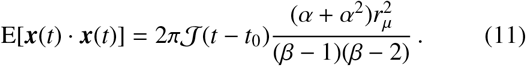

In the left panel of Fig. 5 we have plotted the binned frequency of total daily distances travelled by a population of 5 × 10^4^ individuals following the process defined by Eq. (6) and drawing each individual’s inverse-jump scale 1*/σ* from Eq. (4). We have also fixed *𝒥* = 1, 20 per day (in red and black lines, respectively), *α* = 1 and *β* = 1, 2, 3 as indicated by the increasing opacity within each triplet of lines. Contrasting these distributions, it is immediately clear that by increasing the jump rate *𝒥*, the effect on the distribution of daily distances travelled is similar to increasing the value of *α* — see the middle panel of Fig. 3 for comparison.

**Figure 5.**
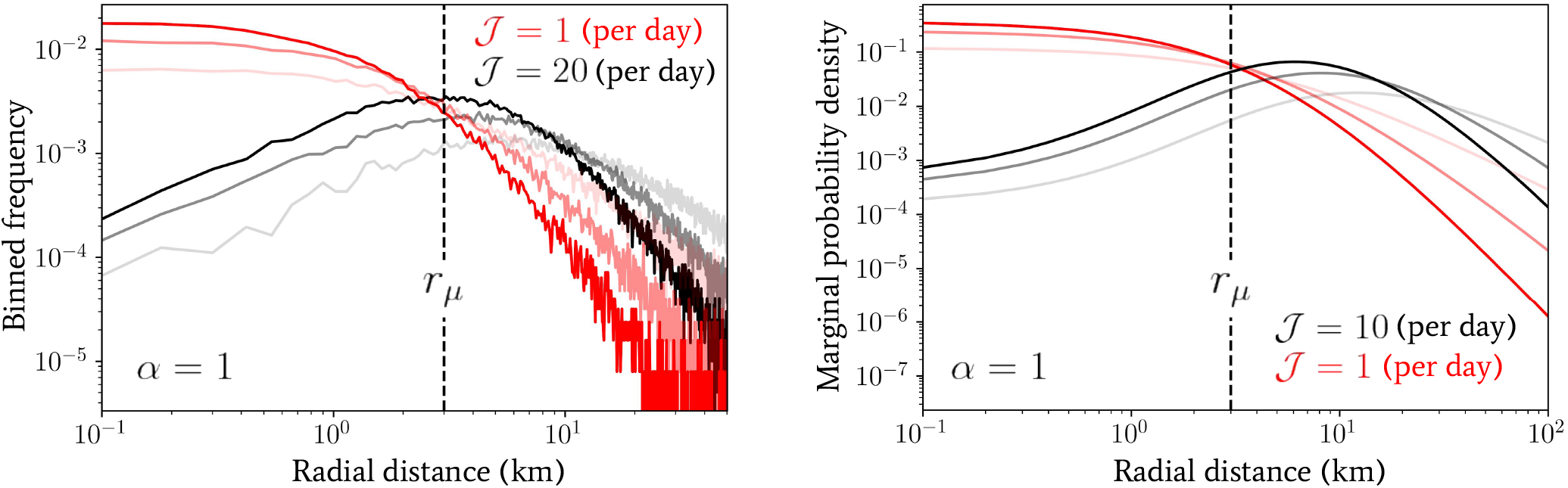
Numerical plots of the distance distributions generated by the multi-jump processes introduced in Sec. 3.1 with randomly-directed (left panel) and in Sec. 3.2 with unidirectional jumps (right panel). In the left panel, a population of 5 ×10^4^ individuals following the process defined by Eq. (6) have been drawn (drawing from Eq. (4) for their jump scale predispositions) and the binned frequency of the total distance evaluated at the end of the day with jump rates *𝒥* of = 1, 20 per day (in red and black lines, respectively), *α* = 1 and *β* = 1, 2, 3 as indicated by the increasing opacity within each triplet of lines. In the right panel, the marginal probability density given by Eq. (15) is depicted with jump rates of *𝒥* = 1, 10 per day (in red and black lines, respectively), *α* = 1 and *β* = 1, 2, 3 as indicated by the increasing opacity within each triplet of lines.

### 3.2. Homogeneous unidirectional jumps

Human movement patterns are not truly random, and instead, one might anticipate a strong directional dependence, e.g., to long distance travel of individuals over the course of multiple days to an intended destination. Due to this fact, let us try to quantify the effect of this behaviour by assuming the logical extreme, i.e., that the angular direction of an individual’s longdistance travel is fixed to a particular value. Following multiple successive jumps while assuming no variation in angle, Eq. (6) then becomes a compound Poisson process for the total jump distance *x*(*t*) ≡ |***x***(*t*) | in a single dimension. This process has a known characteristic function

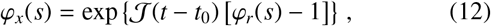

where *ϕ*_*r*_(*s*) denotes the characteristic function of the stationary increments given by Eq. (3). Hence, we may write

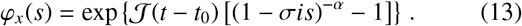

which has no closed-form inverse transformation, but is useful as an expression to calculate the moments. Alternatively, in order to find the distribution over *x*(*t*), we may marginalise over the number of jumps performed (which is Poisson-distributed by construction), which are themselves drawn from the jump distance distribution in the following way

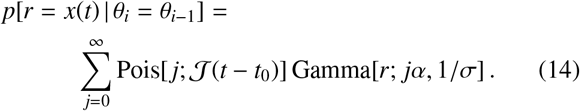

Therefore, if *α* = 1, Eq. (14) yields

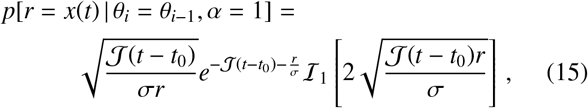

where *ℐ* _*n*_(·) is the modified Bessel function of the first kind.

In the right panel of Fig. 5 we have plotted the marginal probability density given by integrating Eqs. (15) and (4) over 1*/σ* for direct comparison with the one-jump model of Eq. (5). We have used values of *𝒥* = 1, 10 per day (in red and black lines, respectively), *α* = 1 and *β* = 1, 2, 3 as indicated by the increasing opacity within each triplet of lines. Comparing this distribution with the one observed for randomly-directed multiple jumps (see Sec. 3.1), a similar, but even more pronounced, effect by varying *𝒥* is observed — since only a value of *𝒥*= 10 is required to achieve roughly the same change. This is to be expected, as the unidirectional jumpers will always travel at least the same (or more often) a greater distance in total in comparison with the randomly-directed ones, which will likely induce a more severe deformation of the overall distribution when using the former.

The parameter degeneracy between *α* of the one-jump model and *𝒥* in the multi-jump processes described above suggests a test can be performed, to either validate a model, or discard it in favour of a modified version for a given situation with real data. By combining collected data on the total distance travelled per day and the local geometry of distances between buildings travelled to in a given region, the jump rate for each individual (if the same) could be statistically inferred. Comparing this inferred value to real data would provide a test of the movement models we have suggested in this work or potentially provide insight into where they may be improved to better reflect real human daily movement in a given setting.

The simple multi-jump models we have considered in this section have assumed that the distribution for the cumulative number of nearby buildings is homogeneous for each new jump. This approximation is not likely to work well on real-world map data as we have shown there is substantial small-scale heterogeneity in the distances between buildings exhibited in, e.g., Fig. 2. Furthermore, if the multi-jump movement model is purely diffusive over the building locations themselves (itself an assumption which must be investigated further), then there will be correlations in the location distributions between successive jumps that can lead to the distribution of jump distances exhibiting localising effects akin to those exhibited by powerlaw random banded matrices [35]. In other words, when an individual jumps from a cluster of points to, e.g., a point on the edge of their cluster, then they may experience a stronger ‘pull’ back towards the centre of this cluster on the next jump due to the anisotropy in their apparent local point density. This effect should be investigated in future work to evaluate its relevance to movement patterns.

A more sophisticated method to modify the model and deal with (uncorrelated) small-scale heterogeneity could be to sample from the numerically-obtained power laws from Fig. 2 to emulate the spatial heterogeneity in building distances on the real-world map data directly in the processes of Sec. 3. This method would have significant advantages, in terms of computational complexity, to direct simulation methods over the real world map data.

## 4. The effect on reservoirs of infection

### 4.1. A coarse-grained stochastic reservoir network

Up until this point, the movement patterns we have discussed have not been contextualised in an NTD disease model. Since our current (static focal point) framework is best-suited to schistosomiasis and STH transmission, we shall use their well-studied transmission models as a reference throughout this section [36].

Let us now consider a group of focal points that are clustered into a ‘reservoir of infection’. Such a cluster is a source of infectious material from which disease-free individuals are exposed to new infections and, for those already with infections, to potentially increase them in intensity. With each of these clusters, let us now also associate a population of individuals with household locations that are sufficiently close such that the majority of their infectious inputs must be into one of their associated reservoir’s focal points. Note that the vagueness in definition of ‘sufficiently close’ above immediately exposes some of the difficulties associated to defining an EU for the purpose of assessing local prevalence or intensity. Our work in the previous sections of this paper will allow us to proceed with a new approach to quantifying the spatial scales associated with the proximity of households to these focal points of infection. We will then use this approach to understand how spatial scales may be incoporated into the effective dynamical description of STH and schistosomiasis transmission.

Let us now denote the set of indicies which indentify all individuals who live in households that are less than a radial distance *r*_Λ_ away from a focal point that is included in the reservoir cluster as *S* _Λ_. Additionally, to make a reservoir cluster, let us use *r*_Λ_ as a radial separation distance threshold below which any two focal points are clustered together to be part of the same infectious reservoir. For reservoirs of infection of either schistosomiasis or STH, assuming that the associated human population number *N*_Λ_ (the number of elements in *S* _Λ_) does not change, the force of infection (FOI) Λ(*t*) at time *t* is updated according to

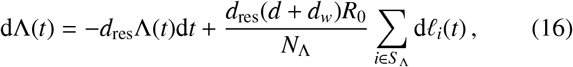

where *d*_res_ is the death rate of the infectious material, *d*_*w*_ is the worm death rate, *d* is the human death rate, *R*_0_ is the basic re-production number and *l*_*i*_(*t*) is a compound Poisson process asociated to the time-dependent infectious material input of the *i*-th individual. Since helminths are dioecious, the definition for *R*_0_, differs from its standard interpretation in standard microparasite diseases since the life cycle of these macroparasites depends on fertilisation of female worms (and hence the presence of both sexes) within a host [36]. Note also that in the equation above, and throughout, we shall neglect age structure in our description — though our formalism can be adapted to include this. This choice is for simplicity in presentation and will not affect our main conclusions.

By clustering focal points into a reservoir of infection by using a radial distance threshold, one might reasonably question the scaling in the spatial extent of the infectious focal points themselves. As was discussed in Sec. 3, the distribution of focal points of infection is likely to be both helminth species-dependent and variable according to local geographic considerations. Due to this variability, the spatial scales associated with clusters of focal points themselves will likely vary across a map. However, this fact will not directly affect the conclusions of the present work since we will consider distances between each individual focal point and its neighbouring households in turn. For specific case studies, the implementation of our algorithm for binding focal points together should be well-defined in most cases for STH and schistosomiasis transmission settings, as long as the distribution of focal points can be inferred. Recall also our previously stated point that, for STH, our household distribution itself could potentially be a tracer for many of the focal points of infection [32, 33].

A more important effect, which is indirectly related to the variability in spatial scale of focal point clusters, is how variable cluster sizes might induce internal variability in the exposure of each individual to new infections. In models of helminth transmission [37], it is most common to account for this variability in exposure through assigning each individual an additional predisposition factor *λ*_*i*_ which is drawn from a gamma distribution, Gamma(*λ*_*i*_; *k, k*), where *k* is the ‘aggregation parameter’ which modifies the variance-to-mean ratio of the distribution of worms within hosts. Due to the epidemiological processes that it aims to capture — and assuming that the basic reproduction number *R*_0_, which accounts for the transmission intensity, remains unchanged — this aggregation parameter will likely vary to capture a change in exposure according to the spatial scale *r*_Λ_ chosen for the reservoir of infection. One simple model to capture this variability in exposure might be to consider the value of *k* itself to be drawn from a gamma distribution, i.e., *p*(*k*) = Gamma(*k*; *a*_*k*_, *b*_*k*_) from which samples are drawn across the map for the individuals closest to each focal point of infection. When binding such focal points together to get the global behaviour of the reservoir of infection, one would then have a mixture model of separate behaviours to consider in the distribution of worms within hosts, affecting the summation term of Eq. (16). Since variability of exposure with spatial scaling is not the main focus of the present work, we shall not investigate this any further here. However, due to its obvious importance to the dynamical behaviour of the reservoir by association, it will be an important component to be cognisent of in future work.

In Ref. [14] it was shown that, to good approximation for moderate-to-large population numbers, the worm burden distributions of individuals *p*(*w*_*i*_, *t*) evolve according to the following reservoir birth-death process master equation

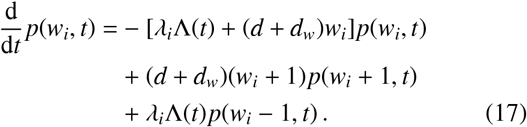

The solution to this equation is a Poisson distribution *p*(*w*_*i*_, *t*) = Pois[*w*_*i*_; *I*_*i*_(*t*)] with time-dependent intensity

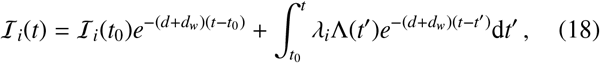

in which we have inserted the solution to Eq. (16). Summing over an ensemble of these individuals to get an overall distribution of worms within hosts, many epidemiogical variables, e.g., the prevalence of infection or mean parasite burden of hosts associated to the reservoir can be calculated (up to specifying the additional gamma-distributed predisposition to infection *λ*_*i*_ for each individual) while maintaining the finite population variance neglected by deterministic disease models. Therefore, in order to understand how these variables which are associated with each reservoir of infection are affected by spatial movements of human hosts, it is essential to correctly assess how Λ(*t*) is modified by inward migration.

Due to the movement patterns we have discussed, there may be individuals who contribute to one of the focal points of the reservoir but come from a region further away such that they are not counted in *S* _Λ_. There may be individuals who are counted in *S* _Λ_ but instead contribute to a focal point outside of their own associated cluster, e.g., on a particular day. Eq. (16) will hence be perturbed by these movements in the following way

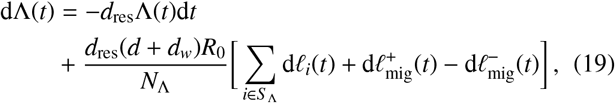

where 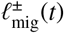 are also compound Poisson processes which sum over the amount of infectious material generated by these migrating individuals that is either net entering (+) or leaving (−) the reservoir focal points.

Note here that Eqs. (16) and (19) currently assume that *N*_Λ_ is static over the time period of interest, which may not be the case if one considers changes that take place over the course of, e.g., a year. This is because long-term effects can cause the population number to vary, including periodic seasonal migrant labour or family visits at particular times of year. Such long-term changes are likely to not have too great a dynamical significance on the terms of Eqs. (16) and (19), however, since these terms are associated to variations in the reservoir of infection which typically occur on much shorter timescales (1*/d*_res_ is typically on the order of weeks or months depending on helminth species).

Eq. (19) implicitly describes a network of spatially coarsegrained reservoirs of infection, each of which varying in spatial extent according to a combination of the distribution of their infectious foci, as well as the spatial coarse-graining scale choice *r*_Λ_. This network of coarse-grained reservoir ‘nodes’ is connected by migratory ‘pulses’, continually affecting each reservoir’s temporal stability (which we shall discuss later on) as well as potentially non-negligible statistical cross-correlations in local epidemiological indicators (e.g., prevalence or intensity of infection) with respect to other reservoirs. Note also that the choice of *r*_Λ_ can therefore control the strength of these migratory interactions between reservoirs in the network. In Fig. 6 we have ilustrated this point explicitly on a digram with real map data based on buildings in central Malawi. The black (red) arrows on the map and zoomed illustrations show migratory movements of a radial distance below (above or equal to) *r*_Λ_, which has been arbitrarily chosen for clear visual effect.

**Figure 6.**
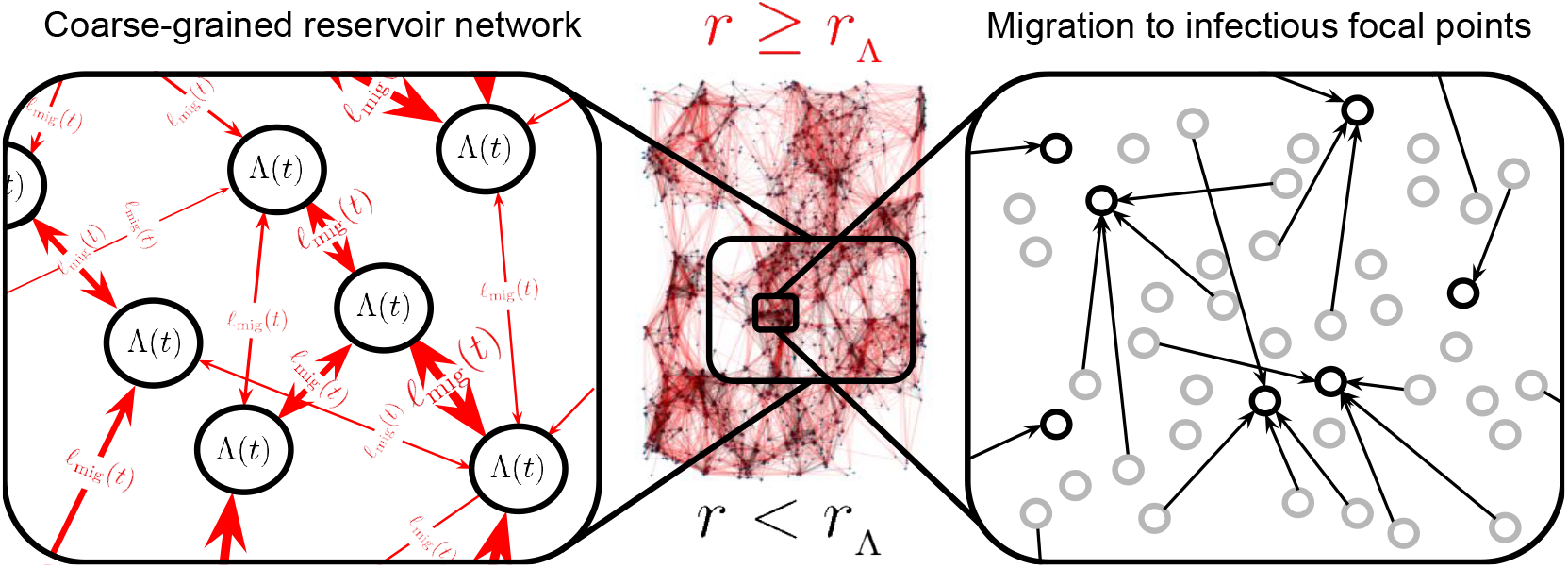
A diagram of real map data (based on buildings in central Malawi) and a zoomed illustration which indicate the coarse-graining procedure of Eq. (21) — in which the black arrows are ‘removed’. Arrows depict individual movements from households (grey hollow dots on the right hand side zoomed illustration) to focal points of infection (black hollow dots on the right hand side zoomed illustration) where black arrows on both the map and its zoomed counterpart correspond to jumps over a radial distance *r < r*_Λ_ (where *r*_Λ_ is the spatial coarse-graining scale) and red arrows correspond to jumps over a radial distance *r*≥ *r*_Λ_. On the left hand side zoomed illustration we see the emergence of the spatially coarse-grained reservoir network ‘nodes’ (nodes are neither drawn to scale nor have geometrically-accurate reservoir spatial shapes) connected by red migratory ‘links’ in time.

Let us now define *T*_Λ_ as the average rate of individuals travel-ling greater than a distance of *r*_Λ_ to a focal point of the reservoir of infection and contributing material to it. Following Ref. [14], this translates into the following decomposition of the positive migratory process in Eq. (19)

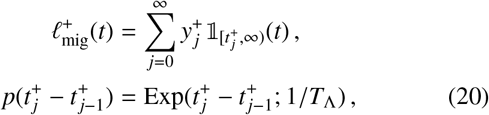

where 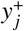 is the contribution to the infectious material quantity of a focal point by an individual: a random variable with a distribution which varies between disease, but likely follows a negative binomial character in some cases. This distribution may be specifically derived by transforming the travelling individual’s worm burden *w*_*i*_ (e.g., drawn from a Poisson distribution and Eq. (18) or an individual-based simulation) into an expected count of fertilised eggs or larvae using a helminth-specific mating function [36]. Note that this compound Poisson process is exact since it tracks the contribution entering the reservoir directly. If one wishes to temporally coarse-grain over the timescale 1*/d*_res_ and see the effect on the mean worm burden deterministic ODE model (as is done in Ref. [13]), an additional non-Markovian component arises to account for the reservoir pulse decay.

When an individual contributes to an infectious focal point which is further than *r*_Λ_ away from their household, they do not just make an additional contribution to the latter’s reservoir of infection, but they also remove their contribution from one of the focal points which is within *r*_Λ_ of their household (their local reservoir). In this sense, each random pulse within the 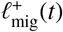 process of one reservoir is exactly correlated to a pulse within the 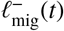 process of another’s. So, for model completeness, it is sufficient to define only the 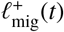 process (attributing an in-In tended location of travel) for each reservoir of infection defined over a map. This is tantamount to defining ‘links’ between the coarse-grained nodes of the network illustrated by Fig. 6.

Let us now consider the influence that the coarse-graining procedure has on Eq. (20). By analogy to physical theories in nature, note that the fundamental dynamical theory should not change — e.g., individual contributions to each reservoir should not ‘disappear’ — simply because of an arbitrary choice for *r*_Λ_, and so one must define a procedure whereby the effective dynamics in Eq. (20) are always describing the same fundamental phenomenon, for any choice of *r*_Λ_. An obvious way to do this is to consider the variation in probability mass associated to individual movement rates as *r*_Λ_ is varied. Based on the arguments of Sec. 2, we hence point out that the following scaling should exist for *T*_Λ_ which accounts for this difference in probability mass arising from a rescaling of distances

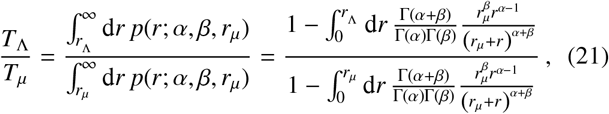

where we have defined *T*_*µ*_ as the average rate of individuals travelling greater than a distance of *r*_*µ*_to a focal point of the reservoir of infection and contributing material to it.

In order to derive Eq. (21) we have assumed that all individuals jump once per day, so that the marginalised jump PDF *p*(*r*; *α, β, r*_*µ*_) of Sec. 2 may be used directly. By integration of Eq. (21), one generally obtains

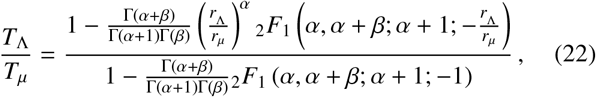

where _2_*F*_1_(·, ·; ·; ·) is the (Gauss) hypergeometric function. Fig. 7 we use the solution given by Eq. (22) to compute the quantity *T*_Λ_*/T*_*µ*_ as a function of *r*_Λ_ */r*_*µ*_ for a range of parameter choices.

**Figure 7.**
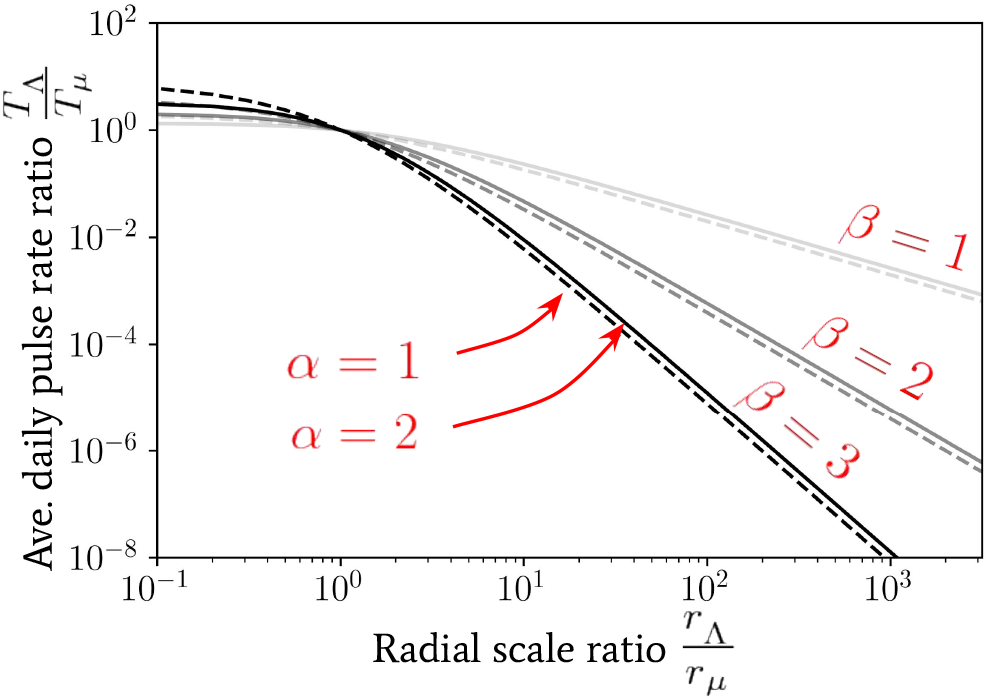
The spatially coarse-grained average daily pulse rate *T*_Λ_ into a focal point of infection as a fraction of its value *T*_*µ*_ from individuals arriving a distance of *r*_*µ*_ or greater away. This value is plotted as a function of the radial coarse-graining scale ratio *r*_Λ_ */r*_*µ*_ used. The relationship is given by Eq. (22) for a range of *α* and *β* power-law parameters.

The variation of *T*_Λ_*/T*_*µ*_ as a function of *r*_Λ_ provides a mapping between the average pulse rate for Eq. (20) that is associated to each focal point within the reservoir of infection (whose pulse amplitudes can be easily summed over to get a total contribution to and from the reservoir) and the distribution of households at different spatial coarse-graining scales. The value of *r*_Λ_ may hence be fixed to identify movement of individuals between structures at different scales, e.g., buildings, villages, towns, cities, etc. We may visualise this coarse-graining of movement distances by referring back to Fig. 6 — there is an emergence of black clusters of connected locations which are connected by red arrows at longer distances. By then clustering the focal points on this map (note this is a mock illustration and these are not real focal points) according to whether or not they are within a radial distance *r*_Λ_ of one another, we can then build coarse-grained reservoirs of infection associated to the scale *r*_Λ_.

It is important to note that infectious reservoir clusters may themselves have a significant spatial extent due to the particular geometry of the nearby focal points, and so *T*_Λ_ should be considered to apply to each and every focal point (or small clusters of focal points) within the reservoir individually. Hence, for reservoirs of infection which correspond to clusters of focal points which, when bound together, have a radial spatial extent that is much smaller than *r*_Λ_ then the results of this section may be used directly for the entire reservoir. In situations where this is not the case — e.g., long and thin geometric relationships binding focal points around the edge of a lake — then our results may still be applied to each focal point (or small cluster of focal points) in turn. The application of our more general formalism is then subject to change depending on the specific relationships between focal points and hence, indirectly, on the species of helminth.

### 4.2. Reservoir Wiener process approximation

Reconsidering Eq. (16), note that the following Gaussian sample mean approximation applies for the terms summing over all (non-migratory) infectious material input into the reservoir

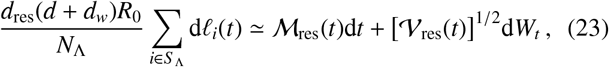

where *W*_*t*_ is a Wiener process, *ℳ* _res_(*t*) is a time-dependent mean and *𝒱* _res_(*t*) is a time-dependent variance for the reservoir inputs. Such an approximation is motivated by the central limit theorem and will be most accurate in the limit of large population number *N*_Λ_. By inserting Eq. (23) into Eq. (19), one finds a drift-diffusion which satisfies the following Gaussian distribution for Λ(*t*)

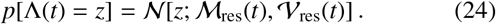

Calculating an exact analytic form for *𝒱*_res_(*t*) in the case of each helminth species is still an open research question since this is very likely to be quite complex to model. For some analytic insight, however, in Ref. [13] it was shown that, with a negative binomial distribution of worms within hosts, the distribution over typical egg outputs from hosts for hookworm is well-approximated by another negative binomial (with different mean and variance). Under these conditions, one may infer, from the sum of negative binomial variances, that

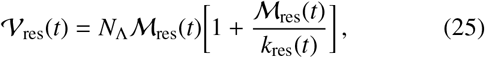

where *k*_res_(*t*) is the aggregation parameter for the *reservoir input* negative binomial, which can be time-dependent before the system relaxes to the endemic steady state configuration.

Eq. (24) provides a significant improvement in computational efficiency over a full stochastic simulation. In particular, for endemic steady-state regions where the constant values of *𝒱*_res_(*t*) and *ℳ* _res_(*t*) can be computed quickly for each parameter configuration, it could be used to generate a simulation likelihood for statistical inference over an entire map affected by helminth transmission and migrating infected individuals.

Due to migratory inputs from (and outputs to) other reservoirs, applying Eq. (24) consistently to each node of our reservoir network will require the inclusion of spatial covariances (or higher-order statistics) between samples. The strength of these spatial covariances can, in principle, be computed by including the migratory terms of Eq. (19) into the approximation above. We can see this by recalling that the rate of these migratory compound Poisson processes (see Eq. (20)) is fundamentally connected to the spatial distance scales via Eq. (22). This ‘fundamentals-based’ method of spatial epidemiological inference for helminth transmission would be distinct from other methods since it would not only include important effects which are inherent to a simulation, such as finite population variance of the underlying stochastic process, but also a theoretical understanding of how the effective dynamical description itself changes with spatial coarse-graining into separate EUs. Note that the ‘scale-invariance’ of the theoretical model we have suggested here can therefore, in principle, be inferred with data collected at one EU scale and then interpolated/extrapolated for model predictions at another.

Although it is beyond the scope of this paper, in future work, it will be of interest to compare the results obtained from other NTD spatial epidemiological models — which typically employ deterministic models a particular spatial scale, e.g., Refs. [9, 10] — with the spatial correlations induced in the diagnostic output from our suggested approach. Such a comparison would help quantify the importance of finite population variance and EU choice on reservoir temporal stability. Other NTD spatial modelling approaches one might consider a comparison to are spatial GRF-based models [6] — these are to be constrasted with the temporally Gaussian, but spatially complex, power-law-like, reservoir network we have proposed in this section. In addition, it would be interesting to explore how the approximate temporal Gaussianity of the model we have proposed in Eq. (24) changes when considering: dynamics further from endemic steady states; configurations closer to transmission breakpoints; or systems with significantly smaller population sizes [14].

### 4.3 The critical spatial scale

In order to evaluate the success of control measures through the calculation of epidemiological observables, such as the prevalence or intensity of infection, it is common for an EU to correspond to a particular spatial scale for the reservoir of infection — see, e.g., [4, 38, 39]. In Sec. 1 we discussed the possibility of defining EUs such that the effect that human movement has on their implied reservoirs of infection was minimal. Note that this is not just of importance to the temporal stability of the reservoirs, but also advantageous because it nullifies any statistical inference bias that may arise through strong migration effects. Such biases may occur when there is a large migration pulse rate of infectious material into a given region where the local intensity and prevalence are increased, but the statistical models using these diagnostics for inference do not include migration rates in their description.

It was shown in Refs. [13, 14] that if the average rate at which reservoir pulses occur exceeds the average death rate of the infectious material in the reservoir, *d*_res_, then the effect of infected human migration on the transmission dynamics becomes particularly strong. In particular, the authors of Ref. [14] demonstrate that, for a range of finite population sizes, migration rates at or above this critical rate into a reservoir of infection can restart transmission in regions where a mass treatment programme has already achieved transmission elimination. In addition, it was shown that this rate of migration can stabilise the stronger ‘fade-out’ effects exhibited near transmission breakpoints. By combining the concept of a critical rate with our proposed spatial model for migration between reservoirs, one may derive a new and important critical spatial scale 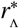 around the focal points of infectious reservoirs which depends on: the type of helminth, the small-scale geometry of locations and the specific patterns of human movement. This spatial scale can be used to define the geographical size of EUs.

If the spatial scale of an EU is chosen which corresponds to a region smaller than the critical scale 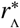 around each of its focal points of infection, the average daily rate of pulses will always be larger in magnitude than the value 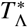, calculated using the critical scale. This is because the curves shown in Fig. 7 are always decreasing with increasing radial scale. This implies that when regions around focal points are defined at this critical scale or smaller for an EU, pulses of the form given by Eq. (19) cannot be safely neglected and should become important to take into account when modelling the transmission dynamics to correctly assess reservoir stability and possible observation biases. This means that if EUs are at this critical scale of smaller NTD programmes need to have measures in place to mitigate the impact of human movement on programme targets. For example, individuals who regularly move beyond the boundaries of EUs should be specifically targeted for treatment. When defining an EU in the opposite case, however, effects from migration between reservoirs of infection may be safely neglected.

Using Eq. (22) the critical spatial scale 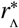 at which 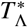 becomes equal to the death rate of the infectious material in the reservoir per day, i.e., 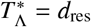, has been plotted for hookworm (which has *d*_res_ ≃ 0.071 [40]) in Fig. 8 as a function of the average daily pulse rate *T*_*µ*_ from individuals travelling distances at or above *r*_*µ*_. In this plot we note that nearly all parameter combinations indicate a sharp decline in the critical scale ratio if *T*_*µ*_ is found to be below *d*_res_ — where, in particular, for values of *α* = 2 the scale ratio appears to fall extremely sharply and hence one can no longer find a finite critical scale below which migration becomes important.

**Figure 8.**
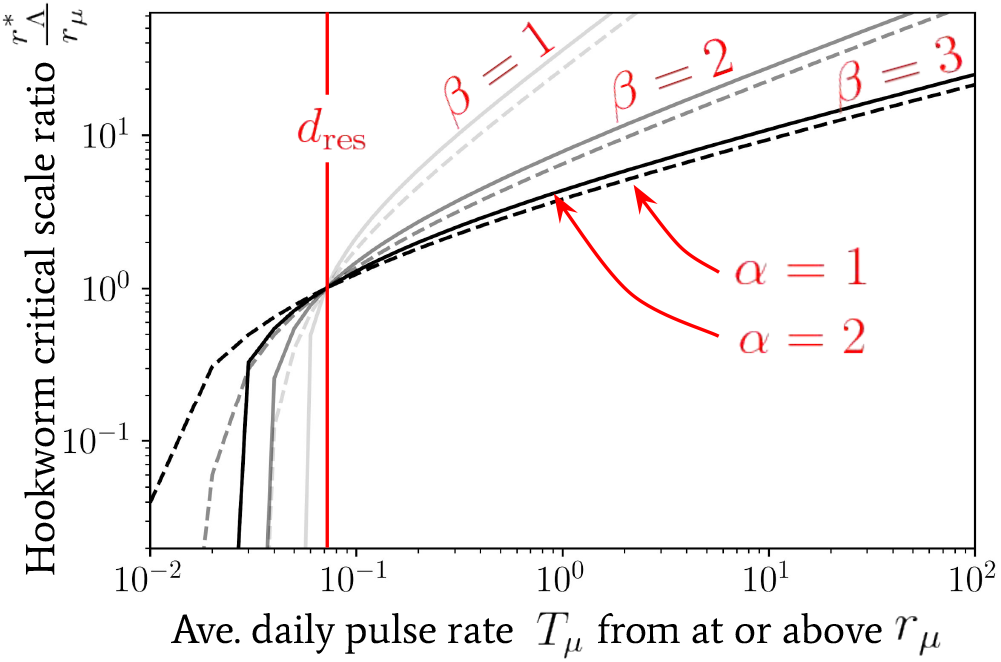
The critical spatial scale as a ratio 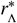 at which 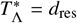, for hook-worm — where *d*_res_ ≃0.071 [40] — plotted as a ratio of *r*_*µ*_. The value of this scale is shown against the value of the average daily pulse rate *T*_*µ*_ at or above *r*_*µ*_. We have used Eq. (22) to generate this relationship with a (bisection) root-finding algorithm.

In contrast, for values *T*_*µ*_ *> d*_res_ of increasing orders of magnitude, one infers from Fig. 8 that the value of *α* is nearly irrelevant (which follows from the construction of *T*_*µ*_) and, particularly for *β* = 2, 3, the increase in the critical scale ratio is very gradual. Such a relationship is also consistent with our expectation as the decline in the average pulse rate from people travelling from distances *r*_Λ_»*r*_*µ*_ is particularly sharp for *β* = 2, 3 — see, e.g., Fig. 7 — and so one requires a significant increase in the amplitude of *T*_*µ*_ to achieve a significant change in the critical scale.

We acknowledge that there are important caveats to the relationship above (and the one shown in Fig. 7) which arise from the assumptions made in obtaining Eq. (21), i.e., the statistical homogeneity and isotropy of people living in the surrounding households who all jump only once per movement event/day directly to an infectious focal point. By relaxing the assumptions made in obtaining Eq. (21), the rate of pulses will vary between different reservoirs of infection. In addition to these modifying effects, the inclusion of multiple unidirectional jumps with different initial preferred directions may no longer be isotropically configured when viewed collectively over the course of the day — which would be the case when multiple individuals all converge to a globally preferred location such as a place of work or a school. We propose to consider such modifications in future work on specific case studies.

Perhaps an even more important model extension to consider was noted in Sec. 4.1, where the potential for the aggregation parameter *k* (for the distribution of worms within hosts) was hypothesised to also vary with spatial scale due to differences in infectious exposure. Although including this potential effect will not directly influence our conclusions with regard to migration, it is possible that another critical spatial scale exists which may be found by considering the value of *k* required to make endemic steady state stability possible. We leave the investigation into the possibility of this additional critical scale to future work.

## 5. Discussion and conclusions

In this work we have developed and studied spatial models of human movement to infectious focal points — the important drivers of new infections for many parasitic organisms in humans. Our movement models have been motivated by the observed patterns of human mobility in, e.g., Refs. [24, 26], but remain robust to setting-specific parametric variations should these observations prove unsuitable for all settings.

At its most basic, our model of human mobility assigns a single daily journey (or ‘jump’) for each individual, however, we have discussed many extensions to this description, which include: multiple may either be in random directions or unidirectional; and substantial local heterogeneity in the distribution of distances between available travel locations. By combining these extensions into a generalised framework, one can generate a wide variety of possible daily human movement behaviours to/between focal points of infection under a unifying description which offers a model-focused explanation for the observed patterns that is also computationally efficient. In future work, it will be interesting to extend our approach to include time dependence in the location of infectious contact events. Such a model will be necessary for diseases such as lymphatic filariasis or onchocerciasis, where bites from mosquito or black fly vectors infected by filarial larvae can vary spatially over time, and may not be Markovian (i.e. memoryless). An investigation into the longer-term temporal changes in population number, e.g., due to seasonal migrant labour patterns or population displacement due to conflict or climate change, may also be interesting.

We note here that the human movement models we have studied are comparable to those developed in Refs. [27, 28] for vectorbourne diseases, which label the single daily journey models as so-called ‘Lagrangian’ mobility models and the multiple successive movement models as ‘Eulerian’. We consider our stochastic approach to generating frequency-distance distributions useful and informative in understanding the underlying probability distributions for these works, especially in relation to demonstrating the regimes in radial spatial scales which are relevant to location-specific geometric effects and human movement distance predispositions.

By clustering focal points into reservoirs of infection, our human movement models have also highlighted the most important information necessary to efficiently (by which we mean low numbers of parameters) describe the effect of migration patterns on these reservoirs. Our model defines a stochastic network of dynamical reservoirs linked by migratory ‘pulses’ (see Eq. (19)) which accounts for these migratory movements at any defined spatial evaluation scale around each focal point using the movement models that we have developed. The necessary data to infer such a ‘reservoir network’ can, in principle, be collected by well-designed field studies. Based on our work here, the information to obtain for a particular study may include:

1. Obtaining the appropriate (likely radially-dependent) power-law *α*(*r*) for the expected cumulative number of locations available to travel to by an individual — see, e.g., Fig. 2. These data can be obtained from geographic maps.
2. Evaluating the amplitude of the effect of small-scale heterogeneity (and perhaps anisotropy) on the power-law cumulative distribution of locations available to travel to by an individual. One can obtain this information from high resolution maps of households.
3. Obtaining the average number of journeys performed within a single day per individual. In this case some metric for having ‘completed’ a journey associated to the time spent at each location will likely need to be determined. In principle, this information can be obtained from questionnaires on human movement that can be integrated into a regular field survey.
4. Evaluating the average number of people entering or leaving the relevant focal points for a given reservoir of infection at some particular spatial scale, e.g., this can be *T*_*µ*_ or evaluated at some other spatial scale since the reasoning of Eq. (21) can be easily adapted. Note that the true spatial scale of the reservoir does not need to be known for collecting these data. This information might be obtained through questionnaires.
5. Potentially obtaining an appropriate scaling value *β* for the distribution of predispositions to jump a certain distance (see Eq. (4)). This may either be consistent with the observed work-home commute patterns in Ref. [24], or deviate to a different power-law relationship entirely, depending on local access to vehicles and other forms of transportation. This information might also be obtained through questionnaires.

While developing our stochastic reservoir network model, we also discussed a computationally-efficient approximate description for its dynamical behaviour near or at endemic steady states. In particular, a Gaussian sample mean approximation was made for the sum of infectious material in each reservoir over a coarse-grained temporal scale, rendering its dynamical behaviour the same as a drift-diffusion process with migratory ‘jumps’ — see Eq. (24). The drastic improvement in computational efficiency that such an approximation offers makes it an attractive option to explore when statistically inferring fully-stochastic infectious reservoirs from diagnostic data. We leave the implementation of this approach, and the analysis of its robustness for reservoir network states far from endemicity, to future work.

The last, and perhaps most significant of our findings in this work concerns the spatial scales associated to EUs. An EU is commonly used to define epidemiological observables such as prevalence or intensity of infection for a given region, often in the context of assessing the perfomance of mass drug administration programmes [21, 20]. By combining our stochastic reservoir network model with a known critical rate for migration into/out of reservoirs of infection, we have been able to demonstrate that a critical radial spatial scale 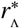 exists around the focal points of infection inside a defined EU. When an EU is spatially defined such that the spatial region of consideration around each of its included focal points is larger than this critical scale, the effects of migration may be safely neglected and our reservoir network may effectively remove its ‘links’. In the opposing limit at scales smaller than 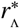 however, we have described how: the temporal stability of the dynamics of infectious reservoirs defined at this scale may be affected through migratory jumps; statistical inference biases may occur through the use of models without migratory effects; and mass drug administration applied only to the population within an EU without considering individuals moving between EUs may experience strong bounce-back effects.

The critical spatial scale we have found in this work is of biological and practical importance to many helminth disease transmission processes. We have also discussed how this model may need further modifications when the relative infectious exposure of individuals is varied according to spatial scale (through the worms-within-hosts aggregation parameter *k*) in addition to many other extensions we have proposed throughout. We therefore consider the model we have presented here to be a useful starting point for many interesting directions for future research.

## Data Availability

Open-source data on building locations are from the high resolution settlement layer dataset generated by the Facebook Connectivity Lab.

https://www.ciesin.columbia.edu/data/hrsl/

## Acknowledgements

RJH, JET and RMA gratefully thank the Bill and Melinda Gates Foundation for research grant support via the DeWorm3 (OPP1129535) award to the Natural History Museum in London (http://www.gatesfoundation.org/). CV grate-fully acknowledges funding from the NTD Modelling Consortium (OPP1184344) by the Bill and Melinda Gates Foundation in partnership with the Task Force for Global Health (http://www.taskforce.org/). The views, opinions, assumptions or any other information set out in this article are solely those of the authors. All authors acknowledge joint Centre funding from the UK Medical Research Council and Department for International Development.

